# Diagnosis of the Beninese Health System: Progress, and Challenges for an Effective, Resilient, and Sustainable System

**DOI:** 10.1101/2024.06.25.24309509

**Authors:** Amboise Tchando Nahini, Mouhamadou Djima Baranon, Emmanuel N’Koé Sambieni, Mouftaou Amadou Sanni

## Abstract

This work involved diagnosing the Beninese health system based on secondary data from several sources (statistical directories, surveys, censuses, and study reports). The descriptive analysis of the evolution of health indicators in Benin between 2002 and 2021 and in-depth interviews with resource people in the health sector made it possible to assess the efforts and progress made by those in power and system stakeholders. There have been many tangible efforts to improve accessibility to health services (the rates of health coverage and attendance at health facilities have increased from 86% to 96% and from 35% to 56% between 2002 and 2021). These efforts have significantly reduced morbidity and mortality rates. The results also indicate a drop in deaths due to malaria (141 deaths to 86 per 100,000 inhabitants between 2003 and 2021) and a drop in the prevalence of HIV AIDS up to 0.8% from 2019 ahead of several countries—neighbors of the sub-region. Early neonatal mortality fell from 10.1‰ to 4.7‰ as well as deaths of children under 5 years old, thus improving life expectancy at birth from 59.6 years in 2002 to 63.8 years in 2013. Despite these efforts, many challenges remain, adding to the strong demographic growth in the country, which clearly expresses the threats weighing on the daily state of health of the Beninese population and calling for a new way of thinking for a sustainable health system. The study ends with the prioritization of challenges and a proposal for strategies to have an efficient and resilient health system, capable of producing quality, healthy, and productive human capital.

## 1 Introduction

The World Health Organization (WHO) defines health as “a state of complete physical, mental and social well-being and not merely the absence of disease or infirmity”[1]. This definition which dates from 1948 and has not been modified until today embraces all dimensions and highlights the importance of good health for development[2]. Thus, the health issue constitutes a major priority for governments and development partners and is included in the golden letter among the 17 Sustainable Development Goals (SDGs) for the 2023 agenda[3]. Several research studies have clearly shown that development and health are inseparable[4, 5, 6, 7, 8, 9, 10]. Many other works explored the issue further by showing the importance of good health for the formation of quality human capital to boost economic growth and contribute to development[11]. Furthermore, the importance of improving health conditions leading to the demographic transition and the economic emergence of many countries has been established and highlighted in several works. This is the case for many Asian countries at the end of the 20th century. In their work where they expose the “miracle of the emergence of the economies of Asian countries”, Bloom and Williamson show how through the demographic transition, these countries once again, at the same level of development as the countries of Africa sub-Saharan Africa around the 1960s, have today become emerging[12]. Later in 2009, Bloom and colleagues outlined the strategies and successes of the emergence of East Asian and North American countries[13]. The significant facts, concrete examples, and practical applications documented at length from the history of these countries, then give glimmers of hope for the emergence of sub-Saharan African countries including Benin.

Furthermore, since 1990, and by its Constitution which guarantees the right to quality health for all, Benin has been resolutely committed to improving the health of its population through several reforms. In 2006, the country launched a new National Health Policy covering the period of 2006-2011 contained in the Strategic Development Orientations Plan, based on the long-term perspectives of “Bénin Alafia 2025”. After evaluation at the end of this policy and given its inadequacies and new health challenges including strong demographic growth, the Beninese State reiterated its actions to promote the health sector by 2030 with a more ambitious and that is consistent with SDG3, which aims to enable everyone to live in good health and promote the well-being of all at all ages: “Benin will have in 2030 a regulated, efficient and resilient health system based on the permanent availability of quality promotional, preventive, curative, rehabilitative and palliative care, equitable and accessible according to the life cycle, at all levels of the health pyramid with the active participation of the population”[14]. The accomplishment of this vision led, especially with the advent of the government, to the disruption of profound reforms, the most important of which aimed to clean up the health sector and improve the quality of health services. These initiatives have certainly led to progress but have not been objectively appreciated over the years and given the resources invested. Likewise, this progress seems apparent because many major challenges remain and weaken the Beninese health system. Strong demographic growth (2.7% per year and an increase of 449% in 62 years) with high rates of urbanization (50% in 2022) constitutes great pressure for the State in covering gaps in health infrastructure and qualified personnel. This situation calls into question the need to rethink the health system differently, which must be seen as a function with several variables, each one as important as the other[15]. Thus, this study seems relevant in the objective assessment of the progress made in the efforts made and all investments in health. This diagnosis, through the analysis of the evolution of Benin’s key health indicators over the years, will make it possible to identify the major challenges of the system to reorient strategic priorities to have an efficient health system that ensures the well-being of the population able to work and contribute to the development of the country.

## 2 Materials and methods

This study was carried out following a mixed approach combining quantitative and qualitative methods. The quantitative approach is more dominant through the analysis of the evolution of key health indicators from several data sources. For its part, the qualitative approach made it possible to deepen the analyses and better explain the quantitative results. They were carried out using qualitative research techniques including observation, interviews with resource people, focus groups, and immersion.

### 2.1 Source of data

This work was mainly based on secondary data from several sources. Data from the Benin Demographic and Health Surveys (DHSB), organized every five years and data from the General Population and Housing Censuses, having a ten-year periodicity as well as population projection data were used. to analyze questions of fertility and demographic growth about the challenges of the health system in Benin. Overall, all of the work was done using data from the statistical directories of the Ministry of Health. These data are produced each year by the Directorate of Planning, Administration and Finance, former Directorate of Foresight and Programming. The health statistics directory provides information on Benin’s health system, including demographic indicators, morbidity, mortality, coverage of health infrastructure, and qualified health professionals. It also provides information on the availability of healthcare services and the epidemiological profile of the country. It gradually integrates community data and that of the private health sector(Ministry of Health, 2022). The data are extracted from the National Health Information and Management System powered by the District Health Information Software 2 (DHIS2), adopted in 2014. We therefore used data from statistical directories available over two decades (2002 to 2021) to analyze the evolution of health indicators to assess the efforts made and identify the weaknesses and main challenges. Apart from the quantitative aspect of the study, great emphasis is placed on the qualitative aspect of this work which affected thirty-eight (38) actors and resource people in the health sector, notably the authorities of the Ministry of Health, leaders of union centers in the health sector, health workers, populations and technical and financial partners involved in health. Using interview guides, four experienced socio-anthropologists conducted in-depth interviews with resource people on the key aspects and challenges of the health sector revealed by the evolution of the indicators in the quantitative part. The themes covered concern health expenditure and human resources, socio-health infrastructure, accessibility and quality of quality services, the health insurance system, and the implications of strong demographic growth.

### 2.2 Analysis methods

This was a descriptive analysis of secondary data. Frequency tables were produced. To make the results easier to read, standard graphs recommended according to the nature of the variables are produced. In addition, we carried out multivariate analyses by crossing several indicators to assess the geographical and socio-demographic differentiation factors. Chi2 independence tests are carried out to assess the significance of the study and possible health disparities. The curve graphs were used to assess the evolution of health indicators between 2002 and 2021. The indicators evaluated concern the different aspects of the health system in particular: health expenditure, human resources in health, health infrastructure and health service attendance, access and quality of health services, and epidemiological profile including morbidity and mortality rates. About the evolution of the indicators from year to year, a critical analysis was carried out of the progress with the explanatory invoices (progress or regression) provided from the results of the interviews with resource persons. These analyses made it possible to highlight the health system’s weaknesses and prioritize the major challenges to be addressed to have an efficient and resilient health system.

## 3 Results and Discussion

### In this section, we present the evolution of health indicators in particular

According to Figure 1, the budget allocated to health has experienced a sawtooth evolution. However, this health budget trend has been declining over time. Although there has been a decline, state allocations to health increased significantly from 2005 to 2010, representing respectively 8.4% and 7.4% of the national budget. The share of more than 10% in 2009 demonstrates the State’s commitment to free cesarean sections. This commitment is evident from the deaths observed during childbirth and the exorbitant cost of cesarean section, which led to the creation in 2009 of the Free Caesarean Section Agency. The latter aimed to improve results in maternal and neonatal health. The agency benefited immediately after its creation from a subsidy of 2.5 billion FCFA in 2010 compared to 0.2 billion FCFA in 2009 and impacted more than 11,543 beneficiaries during the period (2011-2015). From 2011, the drop in the rate increased, going from 6.3% to 4.0% in 2021 and considerable drops have been observed since 2016. These low shares observed are due to the numerous reforms carried out by the government to clean up the sector. We observe a high share in 2020, i.e. 8.8%. This increase in 2020 would be due to the problem of the measures taken by the government due to the response against the Covid-19 disease of which the first case was confirmed on March 16, 2020. In 2021, the State committed 11.54 billion in 2021 to the fight against COVID-19 (ASS, 2021). Table 1 provides information on the essential expenditure items in the public budget devoted to health in Benin.

**Figure 1.**
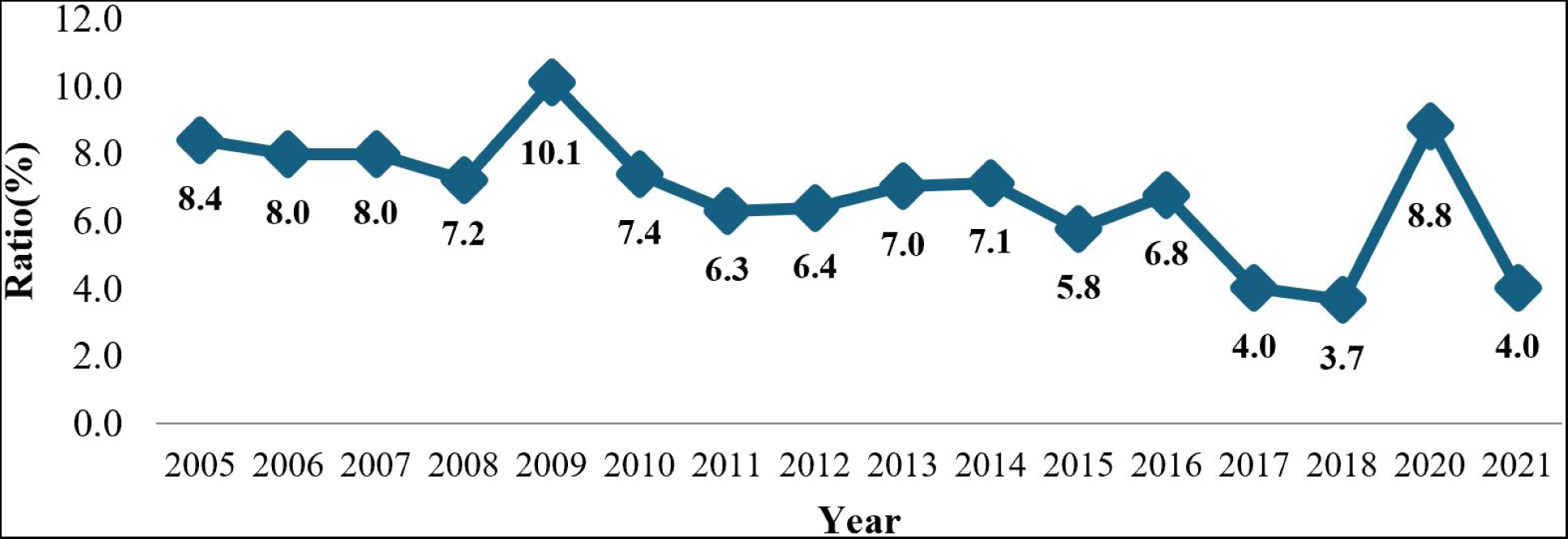
Evolution of the budget ratio allocated to Health as a percentage of the State budget. Source: Calculations from Directorate of Programming and Planning (DDP)/Ministry of Health (Benin) data.

**Table 1.** Budget allocated to health from 2005 to 2021.

| Budget | 2005 | 2006 | 2007 | 2008 | 2009 | 2010 | 2011 | 2012 |
| --- | --- | --- | --- | --- | --- | --- | --- | --- |
| Personnel costs | 7,189 | 8,224 | 8,124 | 6,040 | 24,213 | 18,900 | 12,516 | 13,196.7 |
| Purchases of goods and services | 11,040 | 19,369 | 10,446 | 16,214 | 8,887 | 8,145 | 7,887 | 7,922 |
| Operating transfers and subsidies | 11,244 | 10,144 | 14,321 | 12,070 | 14,746 | 14,746 | 13,737 | 17,737 |
| Socio-Administrative Equipment Budget | 217 | 217 | 295 | 295 | 295 | 1,295 | 1,034 | 1,034 |
| Public Investment Program | 16,765 | 18,963 | 31,695 | 39,302 | 63,194 | 39,377 | 33,980 | 26,846 |
| Total Health Budget (1) | 46,855 | 48,917 | 64,881 | 73,921 | 111,415 | 82,463 | 69,153 | 66,735 |
| General State Budget (2) | 556,923 | 611,216 | 812,561 | 1,023,000 | 1,100,000 | 1,000,439 | 1,099,000 | 1,044,490 |
| Ratio (1) / (2) in % | 8.41 | 8.00 | 7.98 | 7.23 | 10.13 | 7.40 | 6.29 | 6.39 |

| Budget | 2013 | 2014 | 2015 | 2016 | 2017 | 2018 | 2020 | 2021 |
| --- | --- | --- | --- | --- | --- | --- | --- | --- |
| Personnel costs | 19096 | 20854 | 21910 | 23445 | 23138 | 23762 | 24261 | 24489 |
| Purchases of goods and services | 7229 | 7329 | 7934 | 5491 | 5691 | 6312 | 5533 | 7678 |
| Operating transfers and subsidies | 18287 | 19262 | 21120 | 20232 | 20232 | 18185 | 17259 | 22156 |
| Socio-Administrative Equipment Budget | 1004 | 1032 | 1028 | 874 | 874 | 704 | 1000 | 811 |
| Public Investment Program | 27993 | 27838 | 34999 | 19574 | 31131 | 19623 | 78451 | 13475 |
| Total Health Budget (1) | 73608 | 76315 | 86992 | 69616 | 81066 | 68587 | 140274 | 119757 |
| General State Budget (2) | 1044494 | 1069855 | 1506638 | 1026632 | 2010586 | 1862918 | 1592989 | 2985046 |
| Ratio (1) / (2) in % | 7.05 | 7.13 | 5.77 | 6.78 | 4.03 | 3.68 | 8.81 | 4.01 |

This table shows the evolution of the State budget devoted to health and its allocation items. It shows that health budgetary expenditure includes personnel costs, purchases of goods and services, farm transfers, the Socio-Administrative Equipment Budget, and the Public Investment Program. Personnel costs have experienced a roller-coaster ride. Despite this variation, we note that compared to previous years, personnel expenses have doubled in recent years, going from 15% in 2002 to 35% in 2018 and then 20% in 2021. Purchasing costs have seen an increase. generalized decline, going from 24% in 2002 to 7% in 2021. Transfers and operating subsidies generally occupy a quarter of the health budget. As for investment expenses, they account for almost half of the budget of the said sector, representing 36% in 2002 against 44% in 2021. The quality of a health system strongly depends on the human resources that make it up. Let’s now look at the availability of nursing staff in the table below.

**Source:** Calculations from Directorate of Programming and Planning (DDP)/Ministry of Health (Benin) data

Between 2002 and 2021, the number of inhabitants per doctor increased from 7,210 to 13,913, the number of inhabitants per nurse increased from 2,440 to 46,784, and the number of women of childbearing age per midwife increased from 1,555 to 2,301. Significant increases in these figures began to appear in 2018. This increase could be due to the measures taken by the government for the private practice reform which led to the choice of health personnel to practice either in the public or in the private sector. They no longer have the possibility of working in both sectors simultaneously. What about the evolution of accessibility and use of health services?

**Table 2.** Resident/Doctor, Resident/Male nurse, and Woman aged Childbearing/midwife from 2002 to 2021.

| Year | Resident/<br>Doctor | Resident/<br>Male nurse | Woman<br>aged<br>Childbear-<br>ing/midwife |
| --- | --- | --- | --- |
| 2002 | 7,210 | 2,440 | 1,555 |
| 2003 | 7,153 | 3,259 | 1,591 |
| 2004 | 7,135 | 2,648 | 1,705 |
| 2005 | 7,377 | 2,446 | 1,451 |
| 2006 | 7,006 | 1,920 | 1,725 |
| 2007 | 7,472 | 2,045 | 1,510 |
| 2008 | 7,511 | 2,245 | 1,345 |
| 2009 | 7,979 | 2,469 | 1,563 |
| 2010 | 7,979 | 2,469 | 1,563 |
| 2011 | 8,411 | 2,447 | 1,712 |
| 2012 | 5,849 | 1,823 | 1,524 |
| 2013 | 6,418 | 2,006 | 1,645 |
| 2014 | 6,628 | 2,072 | 1,699 |
| 2015 | 6,379 | 2,031 | 1,711 |
| 2016 | 7,014 | 2,169 | 1,898 |
| 2017 | 6,646 | 2,119 | 1,757 |
| 2018 | 23,629 | 4,365 | 2,807 |
| 2019 | 21,137 | 4,441 | 3,302 |
| 2020 | 17,879 | 4,187 | 2,970 |
| 2021 | 13,913 | 46,784 | 2,301 |

**Figure 2.**
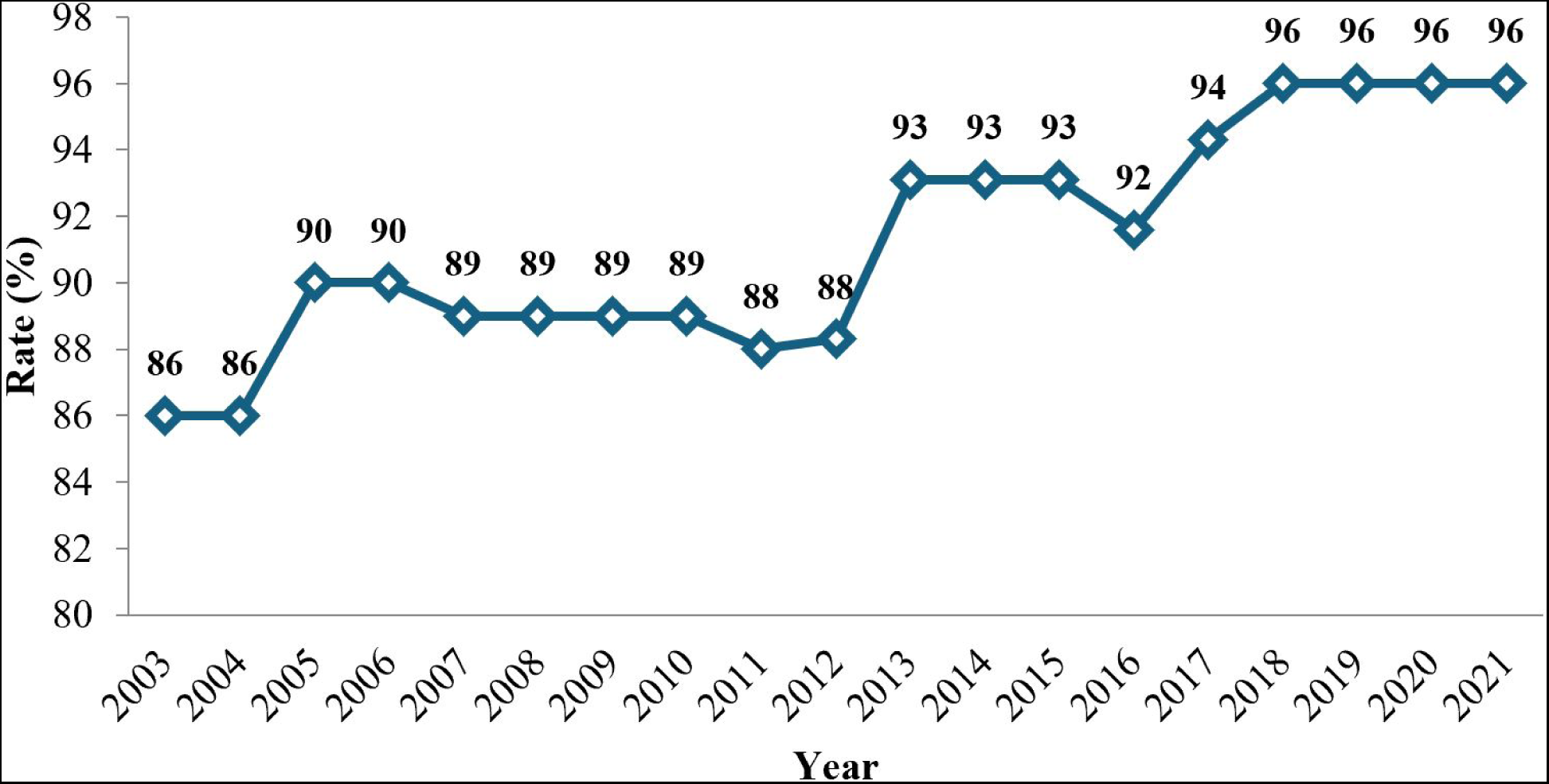
Rate Rate (%) of health coverage between 2003-2021. Source: Calculations from Directorate of Programming and Planning (DDP)/Ministry of Health (Benin) data

The Figure above shows the evolution of the health coverage rate in Benin. According to the analyses, we see an increase in the health coverage rate at the national level, which goes from 86% in 2003 to 96% in 2021. These figures reveal that the country is equipped with health structures capable of caring for the population of the country. However, despite this overall coverage, disparities at the departmental level are observed and deserve attention from health authorities. This would allow all populations the same opportunity to access health services.

**Figure 3.**
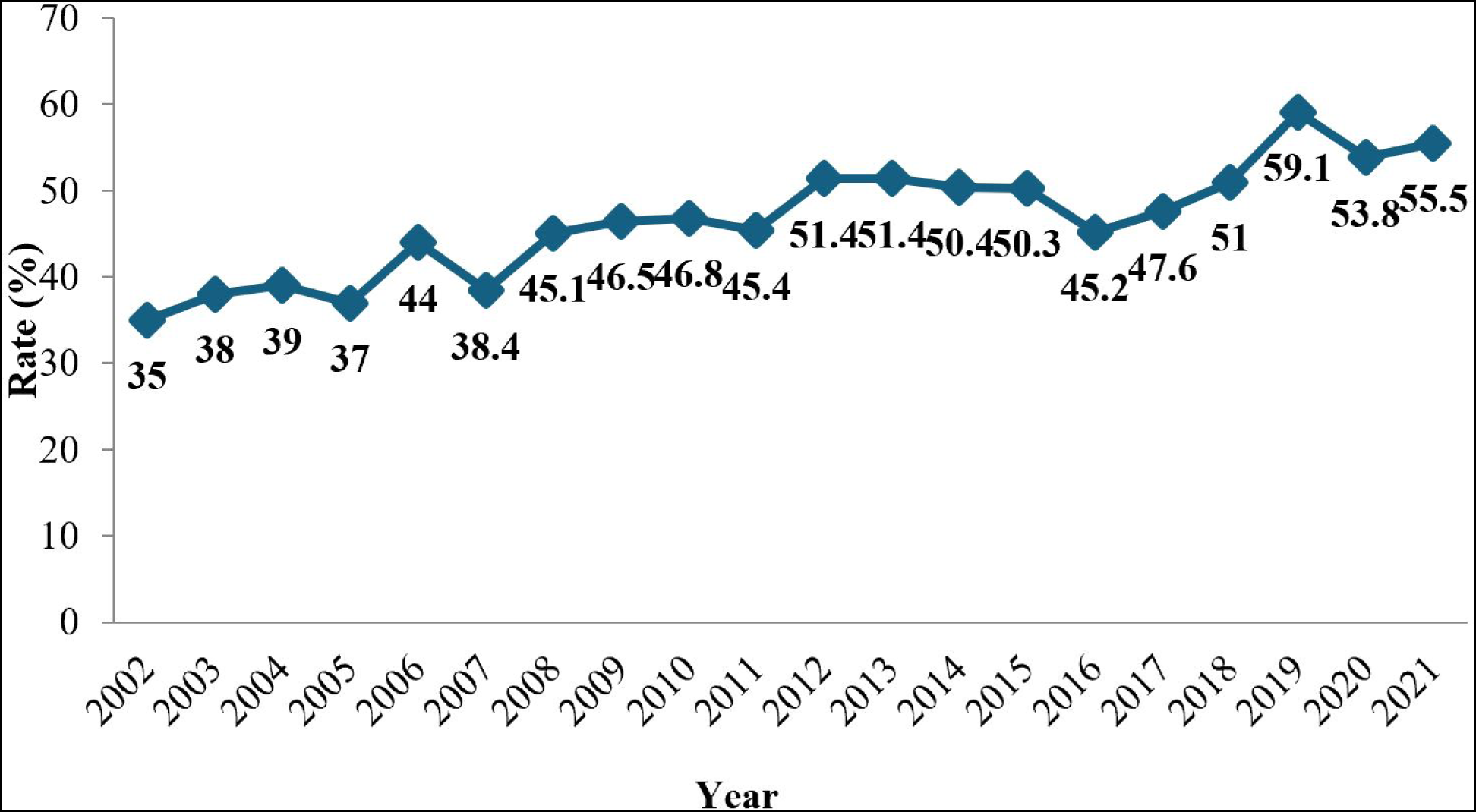
Evolution of the rate (%) of attendance at health services between 2002-2021. Source: Calculations from Directorate of Programming and Planning (DDP)/Ministry of Health (Benin) data

According to this figure, the attendance rate at health facilities is moving up and down. In general, we note a slight change in the attendance of these structures, going from 35% in 2002 to 55.5% in 2021. Nevertheless, the rate of attendance at public health structures by the general population remains average and from one structure to another.

**Figure 4.**
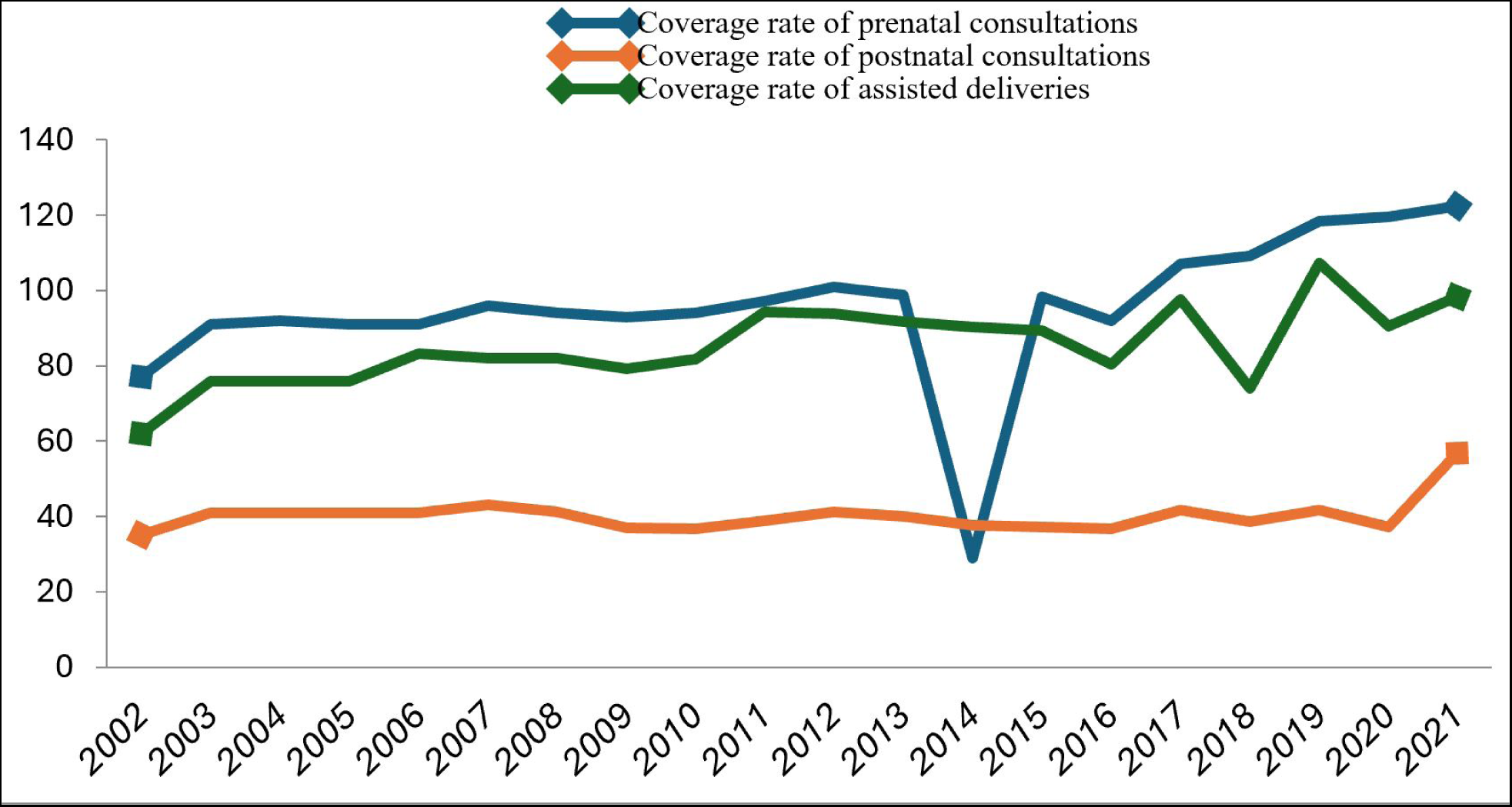
Coverage for prenatal and postnatal consultations and assisted deliveries from 2002 to 2021 Source: Calculations from Directorate of Programming and Planning (DDP)/Ministry of Health (Benin) data

The prenatal consultation rate (ANC) has seen a slight change over time, going from 77% in 2002 to 122.3% in 2021 with only a drop in 2014 (28.9%). Despite this progress, certain indicators raise the issue of super rates (122.3%) and call into question the quality of projections and estimates. Indeed, for these crude indicators (estimated denominators as is the case for the number of pregnancies expected during the year), an underestimation of the denominator leads to super rates and it is difficult in these conditions to objectively assess progress. As for coverage of assisted births, Benin has shown better performance in recent years. Stabilizing around 70% from 2002 to 2005, it rose to 80% from 2006 to 2010 before fluctuating around 90% between 2011 and 2015. From 2016, assistance in childbirth has seen a steady evolution. saw growth increasing from 80.5% in 2016 to 98.3% in 2021. These figures show performance efforts in skilled delivery assistance which can contribute to the reduction of maternal deaths. Regarding postnatal consultations, they are significantly down compared to prenatal consultations, fluctuating between 30 to 40%.

**Figure 5.**
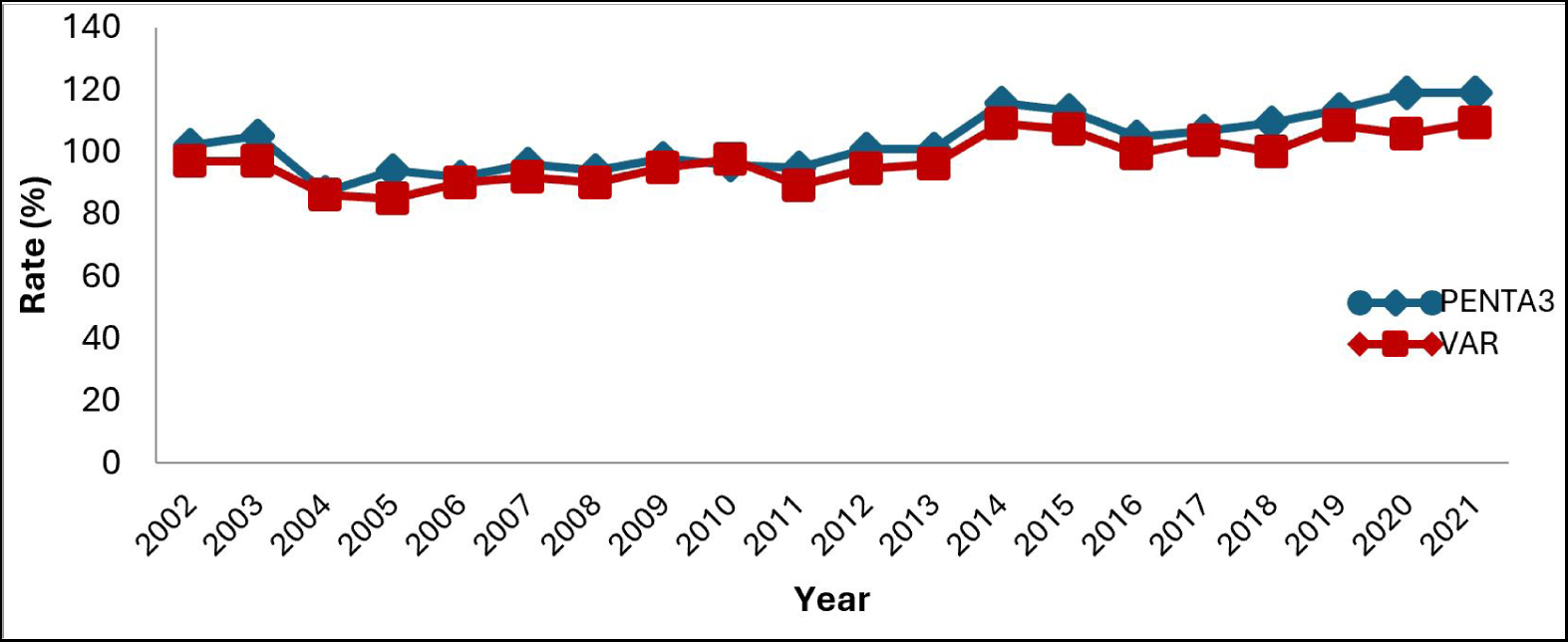
Evolution of penta3 and VAR vaccination coverage from 2002 to 2021Source: Calculations from Directorate of Programming and Planning (DDP)/Ministry of Health (Benin) data

According to this Graph, we see a slight increase over time in the coverage rates for Pentavalent 3 (PENTA 3) and anti-measles (VAR) among children under one year of age. The analysis shows that coverage of the third dose of Pentavalent increased from 102% in 2002 to 119% in 2021, an increase of 17 percentage points. As for VAR coverage, it increased from 97% in 2002 to 109.4% in 2021. These results show efforts undertaken to improve the health of children under 12 months of age.

**Figure 6.**
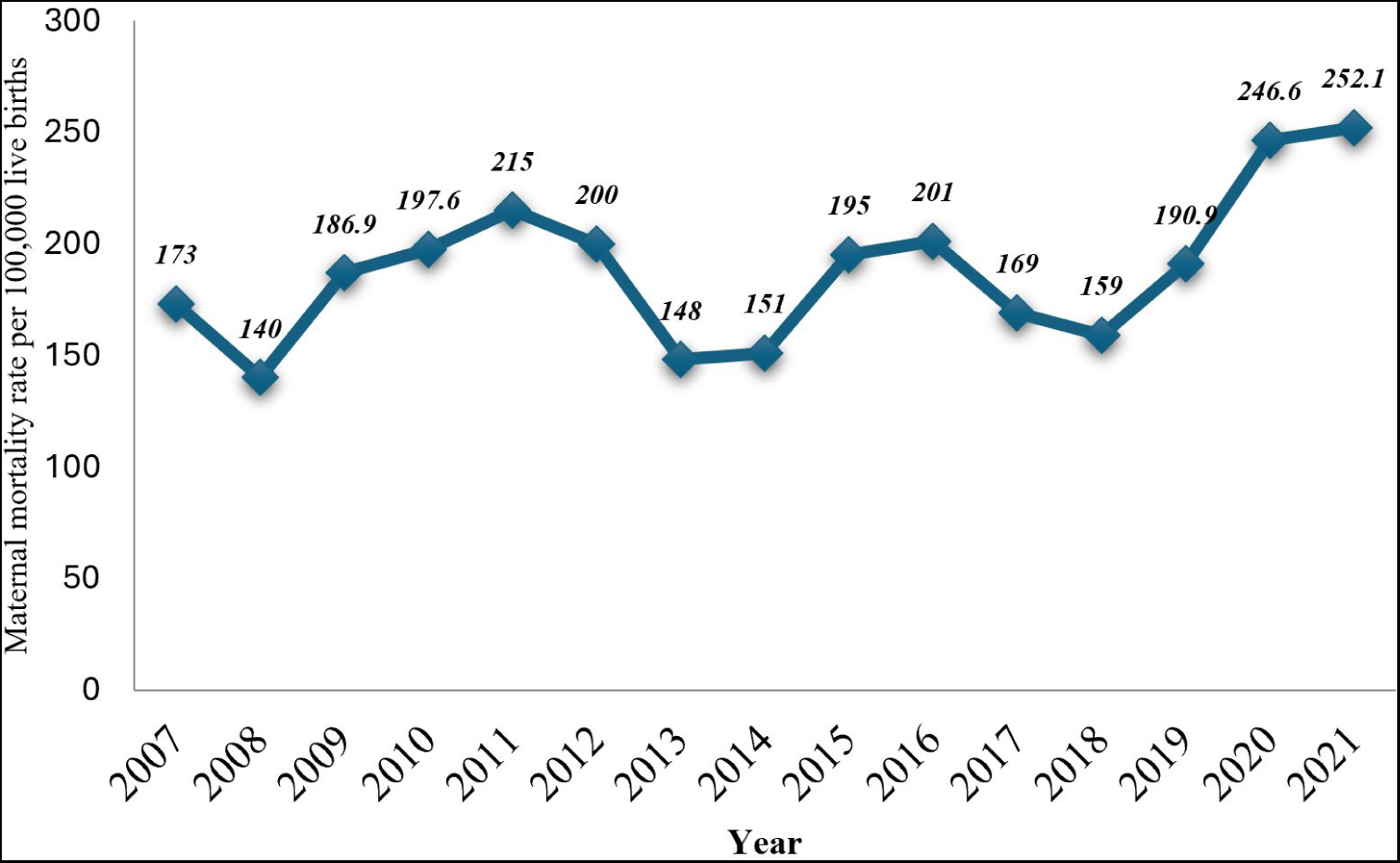
Evolution of the rate of maternal deaths per 100,000 deliveries from 2007-2021 Source: Calculations from Directorate of Programming and Planning (DDP)/Ministry of Health (Benin) data

The maternal mortality rate per 100,000 hospital deliveries changes with some fluctuations over the years. It went from 173 per 100,000 deliveries in 2007 to 215 in 2011. It gradually decreased and reached 148 per 100,000 deliveries in 2013 before experiencing an increase to 201 in 2017. This rate decreased to 159 per 100,000 deliveries in 2018 before to rise to 252.1 per 100,000 deliveries in 2021. This trend in deaths in health centers confirms the insufficiency of the effort made to reduce maternal deaths. There is an urgent need for greater commitment to reduce maternal mortality to achieve the SDGs.

**Figure 7.**
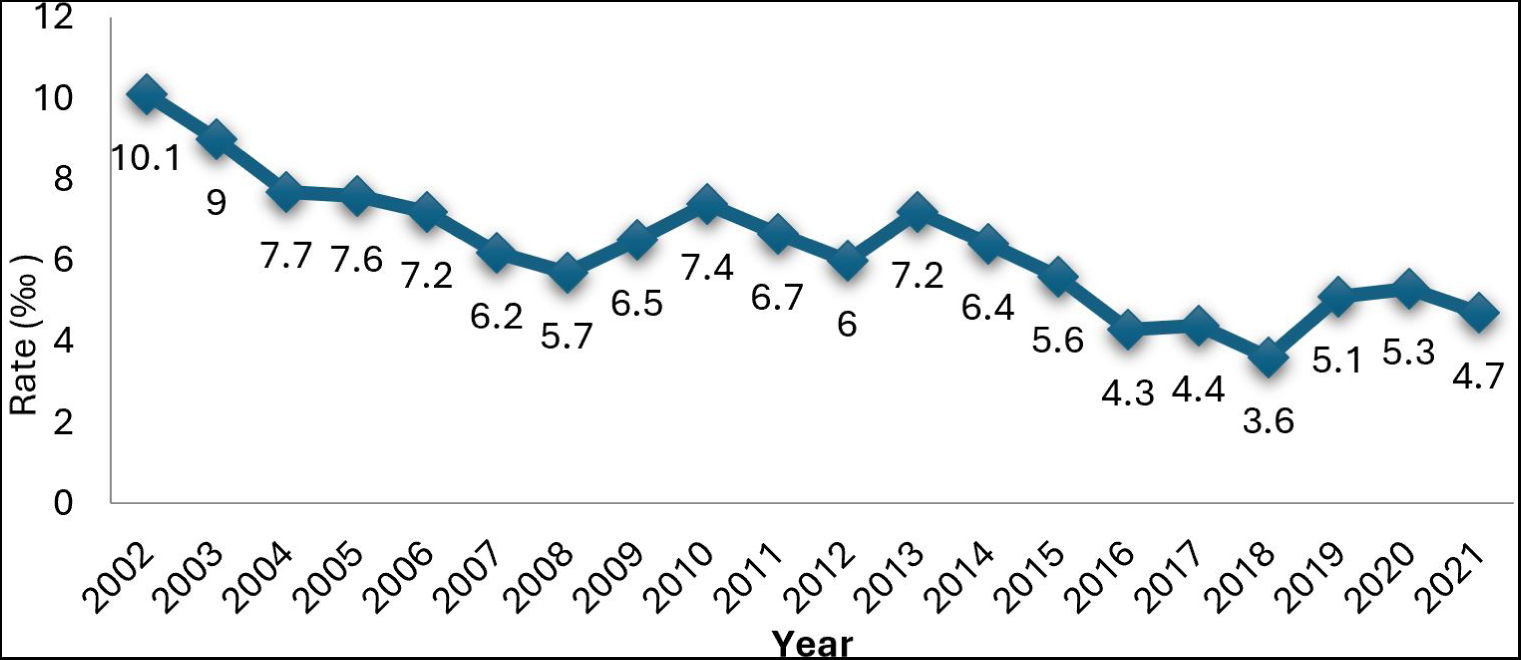
Rate (‰) of early neonatal mortality by department from 2020 to 2021 Source: Calculations from Directorate of Programming and Planning (DDP)/Ministry of Health (Benin) data

In Benin, early neonatal mortality decreased from 10.1‰ in 2002 to 4.7‰ in 2021. However, this decline remains insufficient because neonatal mortality has been identified as that which contributes to infant deaths. Efforts must be made to adequately equip hospitals with equipment and qualified personnel for the care of newborns. Furthermore, early neonatal mortality also experiences disparities by department. The graph below shows this disparity.

**Figure 8.**
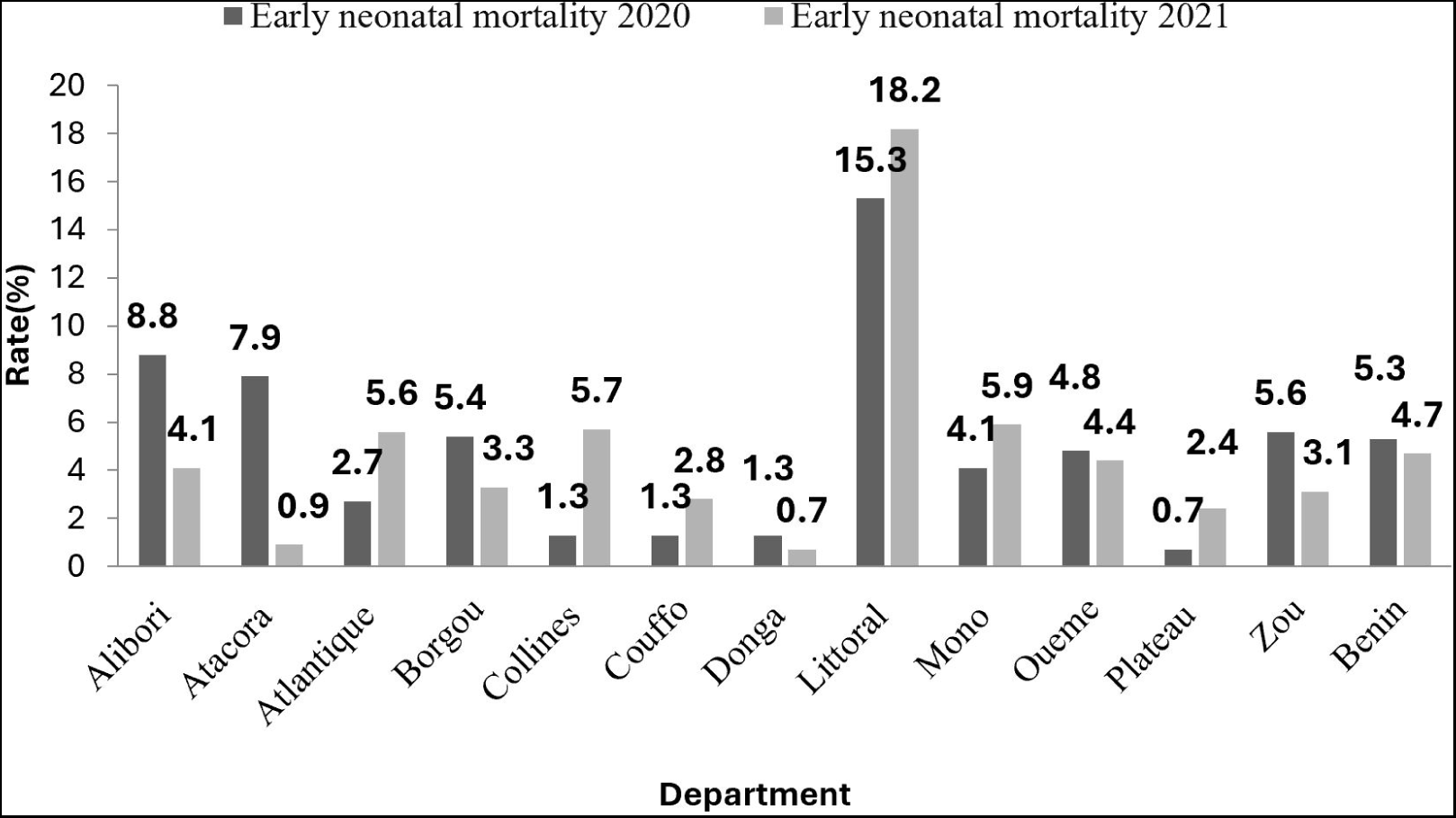
Early neonatal mortality rate by department Source: Calculations from Directorate of Programming and Planning (DDP)/Ministry of Health (Benin) data

Between 2020 and 2021, although early neonatal mortality is stabilized around the overall rate of 5.3‰ and 4.7‰ respectively, the Littoral displays a very high neonatal mortality which increased from 15.3‰ in 2020 to 18 ‰ in 2021. On the other hand, the departments of Donga, Plateau, and Couffo have the lowest rates due to early neonatal mortality every two years combined, representing 1.3‰ compared to 0.7‰, 0.7‰ against 2.4 and 1.3‰ against 2.8‰. Efforts still need to be made to reduce this rate within each department. The one in Cotonou must pay particular attention. Furthermore, most child deaths are due to malaria. The following graph shows the evolution over time of the malaria incidence rate.

**Figure 9.**
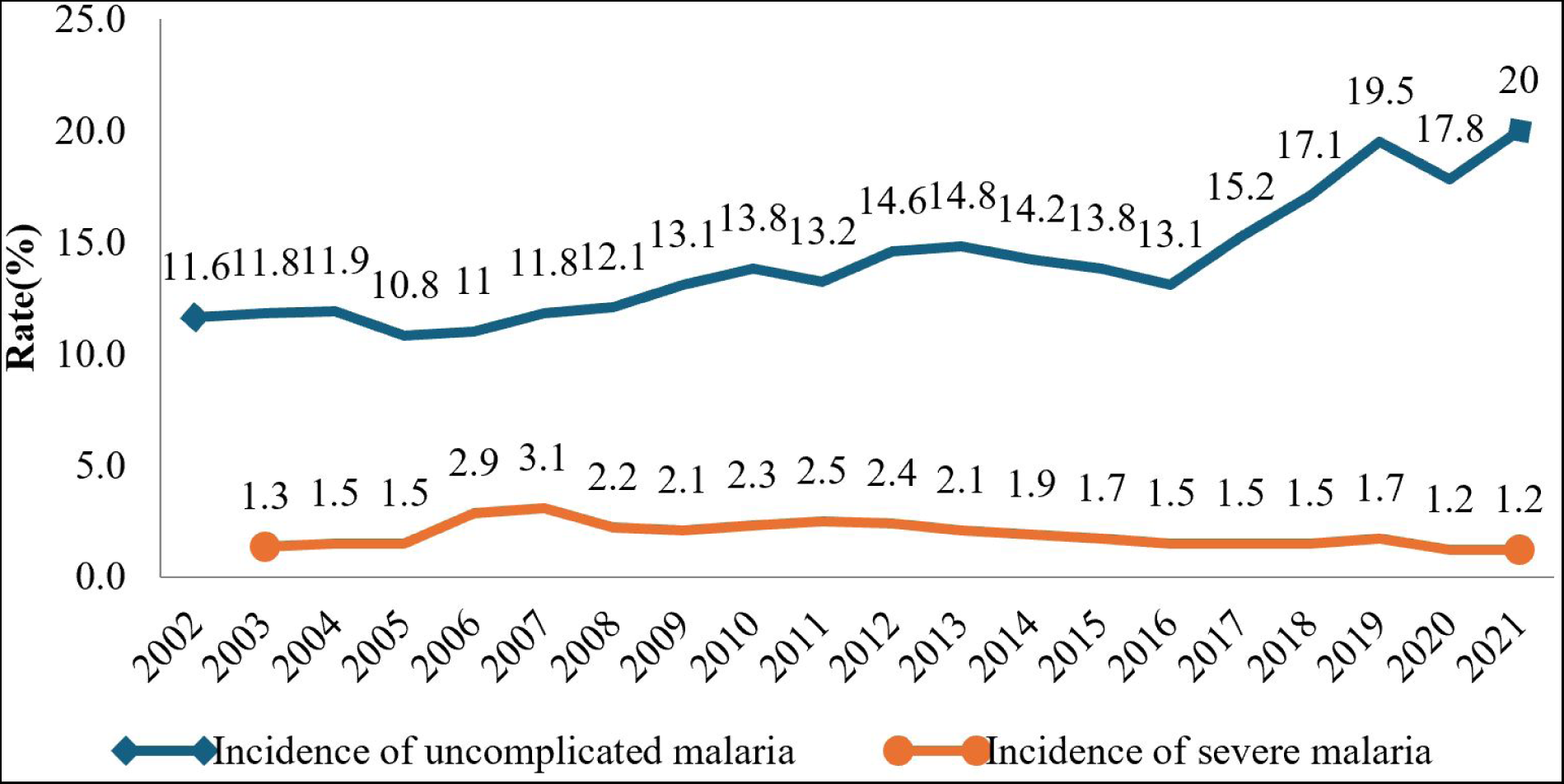
Incidence rate (%) of uncomplicated and severe malaria Source: Calculations from Directorate of Programming and Planning (DDP)/Ministry of Health (Benin) data

Designating the major public health problem in Benin and the leading cause of hospitalization for the population, particularly for children under 5 years old, i.e. respectively 18% and 29% of cases, the incidence of malaria continues to grow over time. The malaria incidence rate is the ratio of the number of malaria cases recorded per 1000 inhabitants. Mainly, the analyzes of this graph shows that the incidence rate of uncomplicated malaria in Benin is growing sharply from 11.6% in 2002 to 20% in 2021. This shows that efforts still need to be made to eradicate the incidence of malaria in Benin. On the other hand, the incidence rate of severe malaria has declined in recent years and stabilized around 1% in 2021. Severe malaria results from delayed treatment of uncomplicated malaria and easily leads to death. This progress in the control of serious malaria could be explained by the implementation of community health through basic care offers that reduce the occurrence of serious cases.

**Figure 10.**
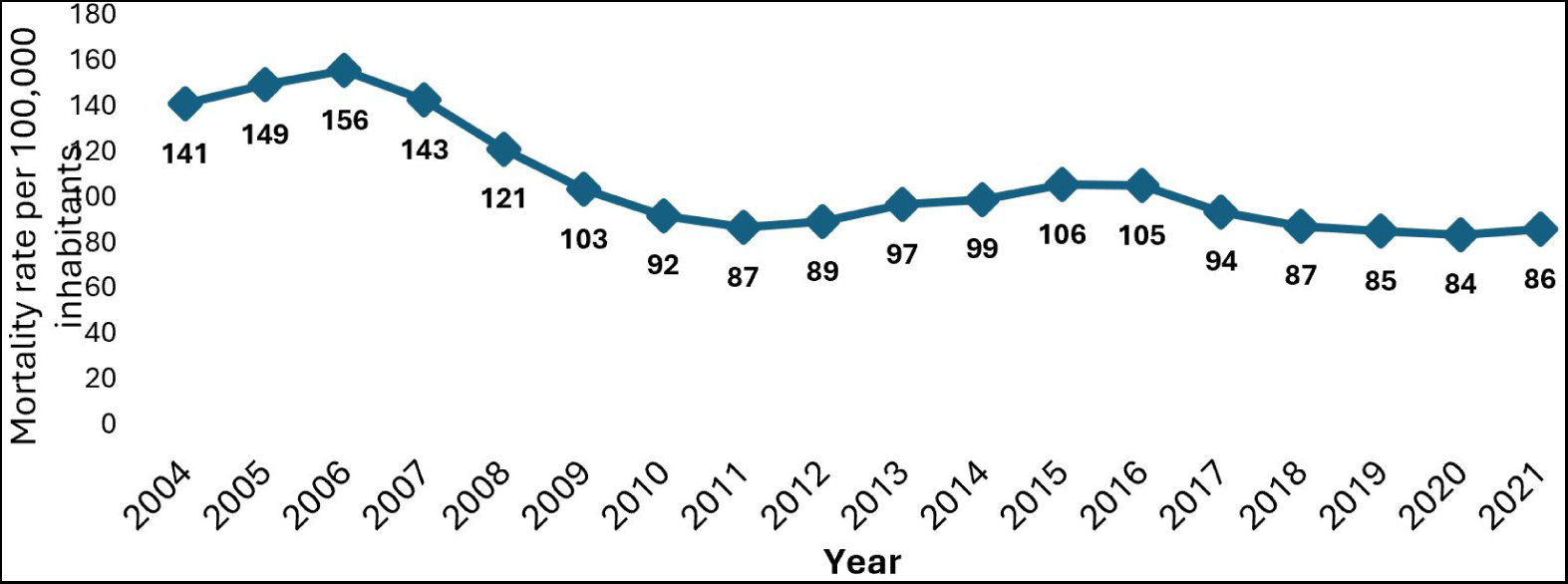
Malaria mortality rate per 100,000 inhabitants Source: World Health Organization (WHO)

Deaths from malaria have seen a significant decline in recent years. From 141 in 2004 to a peak of 156 per 100,000 inhabitants in 2006, the mortality rate due to malaria experienced a gradual decline before stabilizing around 80 per 100,000 inhabitants in recent years. This decline may be associated with the control of severe malaria and efforts to cover the territory with insecticide-treated mosquito nets (ITN). The graphs below show the evolution of ownership and access to impregnated mosquito nets. by households.

**Figure 11.**
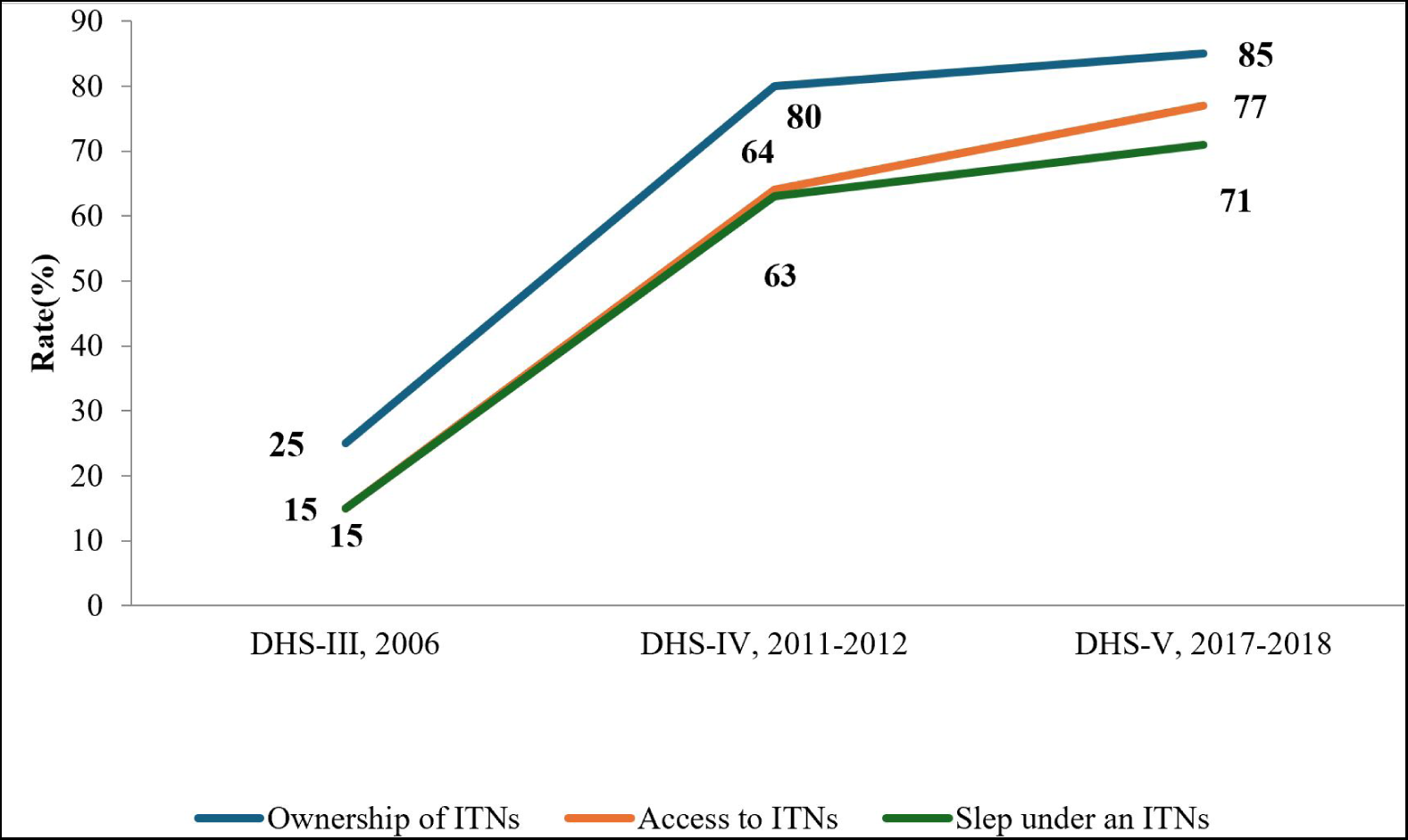
Percentage of households owning, having access to, and having slept under mosquito nets an ITN Source: INSTaD: Demographic and Health Survey (DHS) Benin III, IV and V

The State has engaged, with the support of these partners, in the distribution of impregnated mosquito nets in recent years. Analyzes reveal a high percentage of households owning mosquito nets, increasing from 25% in 2006 (Demographic and Health Survey (DHS) Benin-III, 2006) to 85% in 2017-2018 (Demographic and Health Survey (DHS) Benin -VI, 2017-2018). According to the latest demographic health survey, a household in Benin has an average of 2.3 mosquito nets impregnated with insecticide. The population sleeps under mosquito nets as much as they have access to them. In 2006, 15% of households had access to ITNs and also, 15% used them. This proportion increased from 77% and 71% in 2018, a difference of 6%. This gap shows that not all households with access to mosquito nets use them. This can be explained by the fact that some people use mosquito nets for other purposes, notably for bird cages, fences, gardens, etc. HIV/AIDS is one of the priority diseases and is the subject of a lot of attention by leaders with the support of partners. He would be interested in assessing the evolution of the prevalence of HIV/AIDS in Benin.

**Figure 12.**
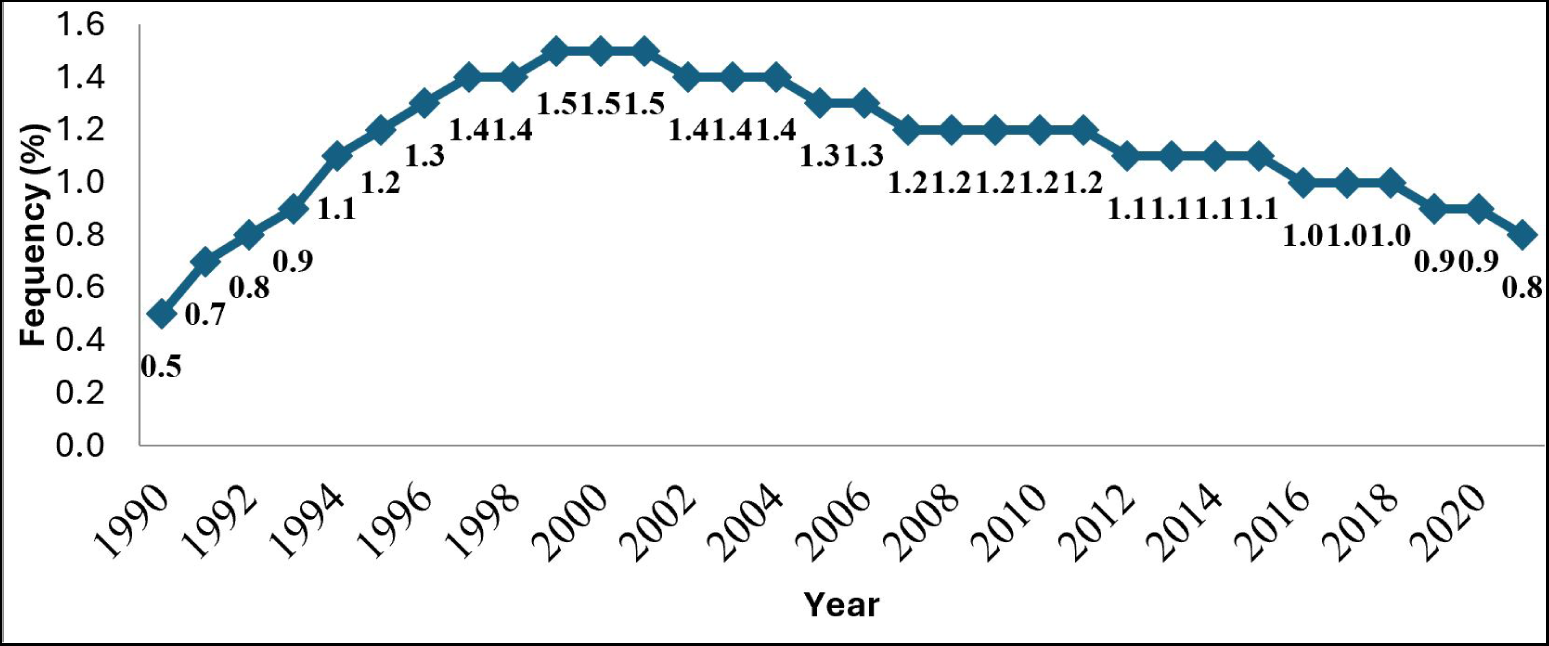
HIV prevalence as a percentage of the population aged 15-49 Source: World Health Organization

In Benin, HIV prevalence among people aged 15 to 49 reached a peak of 1.5% between 2000 and 2002 before falling significantly in recent years. After stabilizing around 1.2% between 2008-2011, it fell by a value of less than 1% from 2019. This shows the State’s commitment in the prevention of this disease within the population. The Health Program to Fight AIDS therefore uses good strategies with the support of these partners to combat HIV/AIDS.

**Figure 13.**
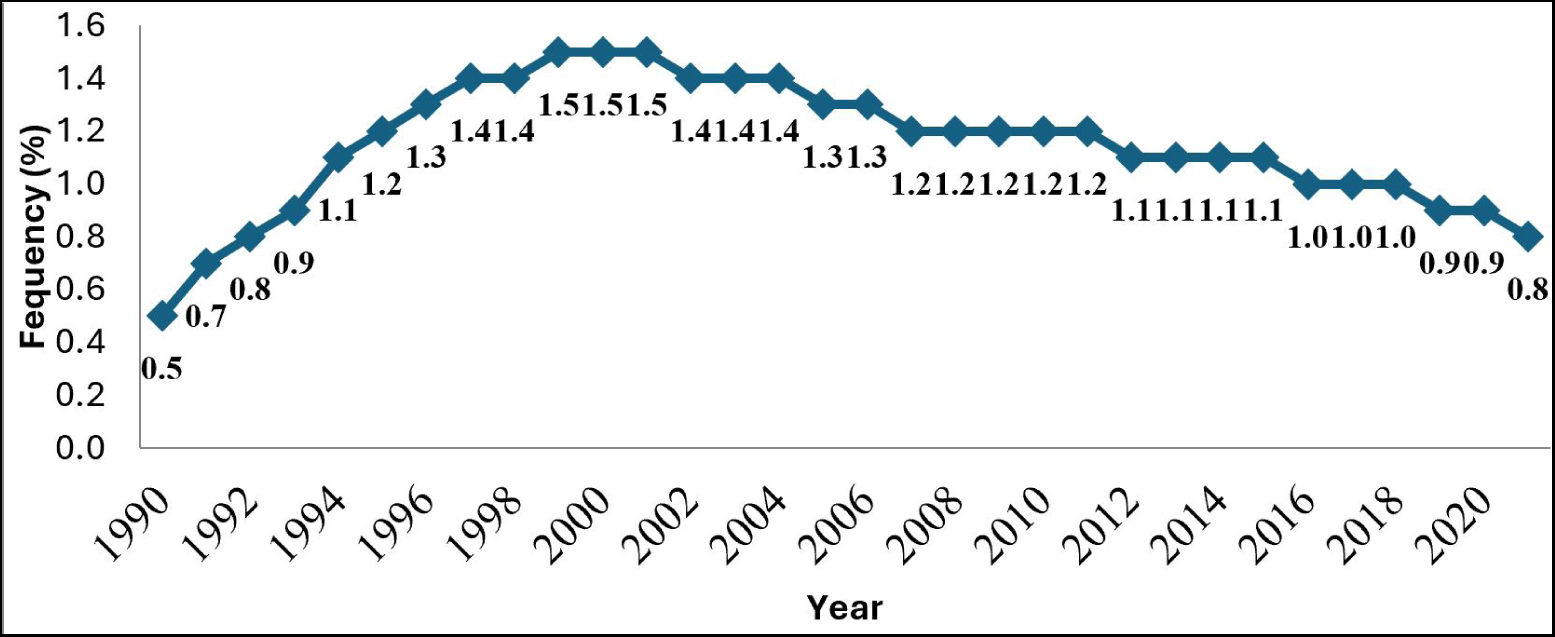
HIV prevalence by age and gender in 2011-2012 Source: INSTaD: Demographic and Health Survey (DHS) Benin -IV 2011-2012

In 2011, when HIV prevalence was 1.2%, there was a difference by age and sex. Generally at all ages except in the 15-19 age group where HIV prevalence is low, i.e. 0.1% compared to 0.6% in men, and at 45-49 years where there is no gap (1.3% each), women are more exposed to HIV/AIDS than men. However, it is adults aged 30-34 who are more exposed to the risk of HIV with a seropositivity of 2.4% among women compared to 1.6% among men.

**Figure 14.**
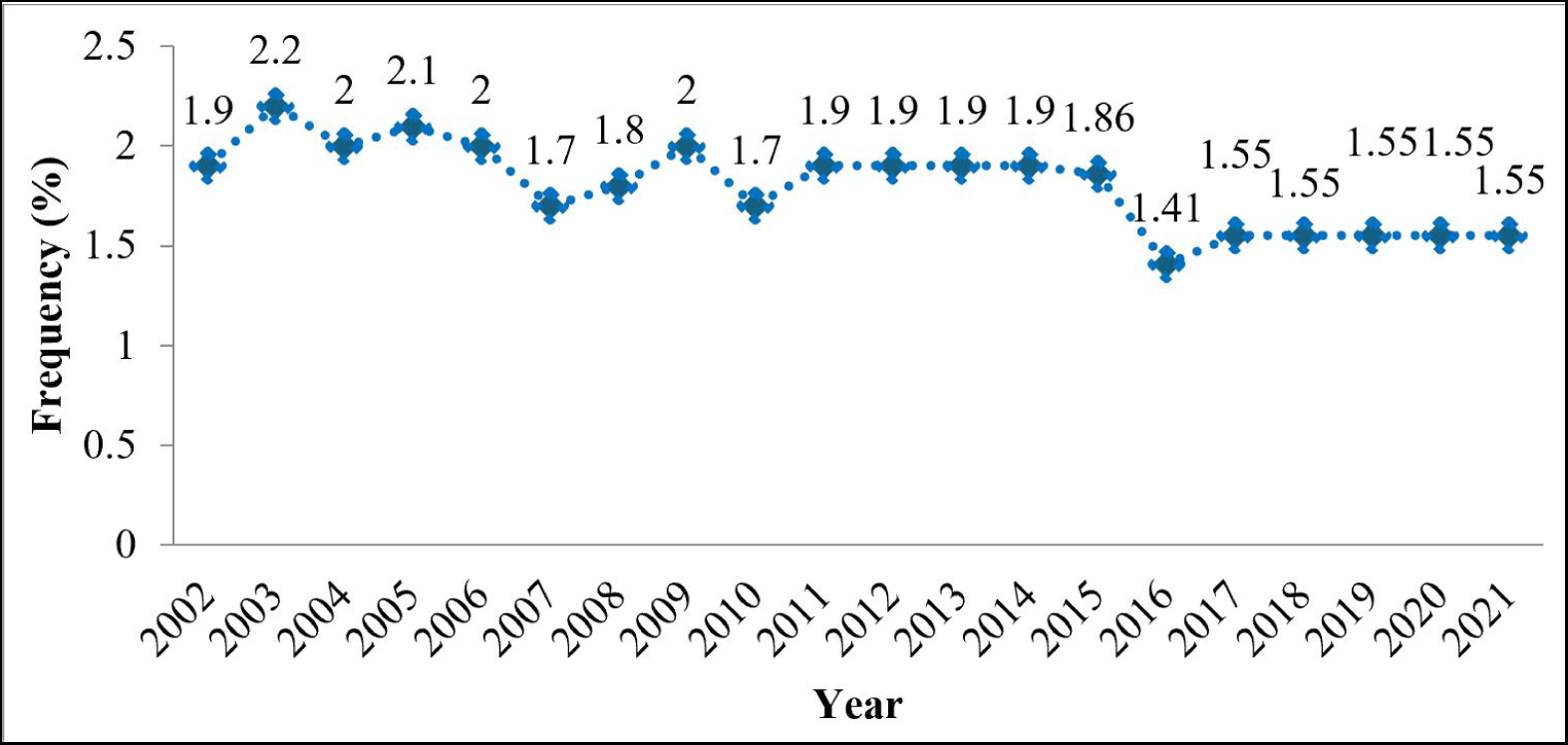
Prevalence (%) due to HIV/AIDS among pregnant women between 2002-2021 Source: Calculations from Directorate of Programming and Planning (DDP)/Ministry of Health (Benin) data

The prevalence of HIV/AIDS among pregnant women has seen an ups and downs pattern with some stabilization in recent years. From a prevalence of 1.9% in 2002 to a peak of 2.2% in 2003, HIV prevalence fluctuated around 2% between 2004-2009, stabilizing at 1.9% from 2011 to 2015 before returning to 1.55% from 2017. These efforts demonstrate the quality of control of this disease among pregnant women. However, disparities remained observable between the country’s departments. The table below reveals the differences in HIV prevalence among the population of pregnant women.

**Table 3.** Prevalence of HIV among pregnant women by department in 2017.

| Department | Urban | Rural | Total | 95% CI |
| --- | --- | --- | --- | --- |
| Alibori | 0.4 | 0.57 | 0.51 | 0.40-0.61 |
| Atacora | 1.81 | 0 | 0.68 | 0.58-0.78 |
| Atlantic | 1.58 | 0.33 | 0.83 | 0.71-0.94 |
| Borgou | 1.72 | 0 | 0.68 | 0.58-0.78 |
| Collines | 1.59 | 0.9 | 1.17 | 1.02-1.32 |
| Couffo | 4.29 | 1.29 | 2.36 | 2.16-2.56 |
| Donga | 2.65 | 1.18 | 1.69 | 1.51-1.86 |
| Littoral | 1.64 | - | 1.64 | 1.39-1.89 |
| Mono | 2.09 | 0.39 | 0.98 | 0.85-1.11 |
| Ouémé | 2.81 | 1.58 | 2.06 | 1.87-2.26 |
| Plateau | 2.01 | 1.42 | 1.63 | 1.45-1.81 |
| Zou | 1 | 0.85 | 0.91 | 0.78-1.05 |
| Benin | 1.97 | 0.77 | 1.55 | 1.31-1.79 |
|  | (1.70-2.24) | (0.60-0.95) |  |  |

In Benin where the prevalence of HIV among pregnant women is 1.55%, the departments of Couffo and Oémé are those where it is higher, representing respectively 2.36% and 2.06%. Efforts must still be aimed at these departments in order to lower this rate. On the other hand, the prevalence among these women is below 1% in Alibori (0.51%), Atacora (0.68%), Borgou (0.68%), Atlantique (0.83%), Zou (0.91%) and Mono (0.98%).

**Figure 15.**
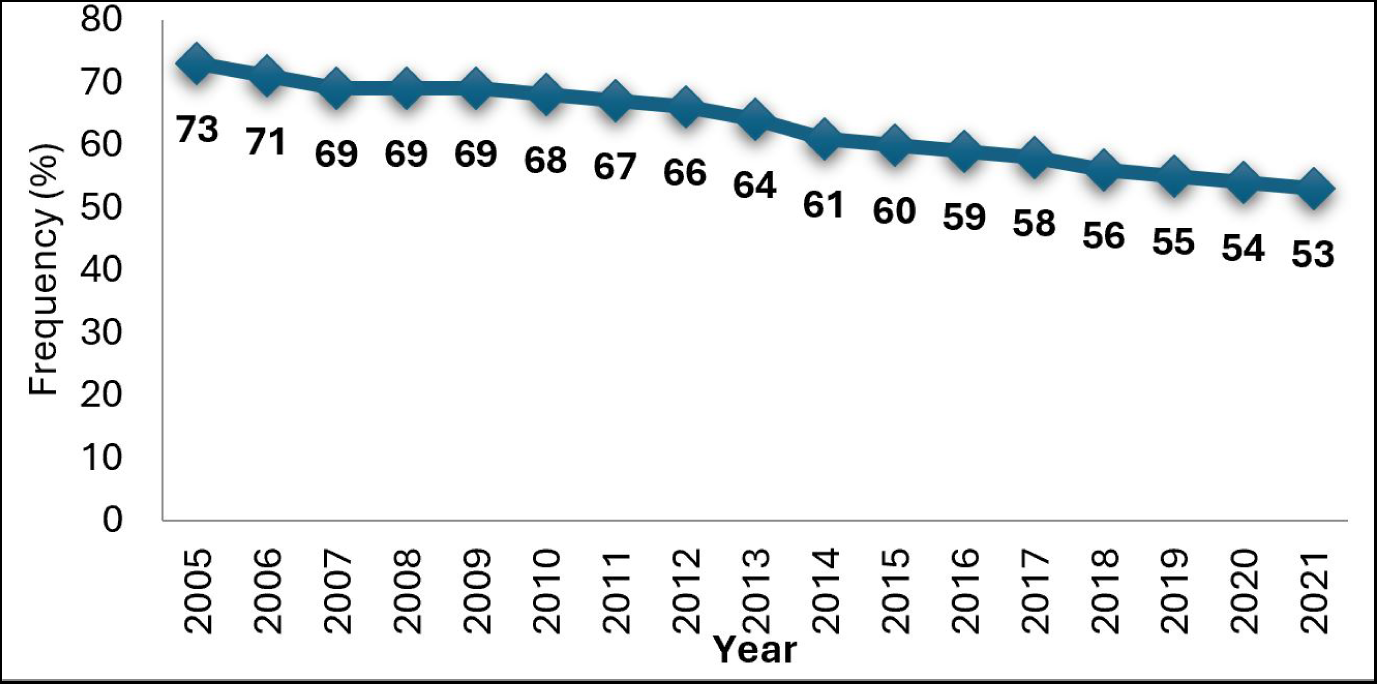
Incidence of tuberculosis per 100,000 inhabitants Source: Calculation based on World Bank data

In Benin, the incidence of tuberculosis is continuously decreasing, from 73 in 2005 to 53 per 100,000 inhabitants in 2021. Efforts to combat this disease have borne fruit. One of the strategies adopted by the Ministry of Health to prevent maternal and child deaths is family planning through promoting the adoption of modern contraceptive methods. Numerous means are being put into making contract methods accessible to populations, particularly women of childbearing age. Do contraceptive prevalence rates follow the efforts made and the resources invested?

**Figure 16.**
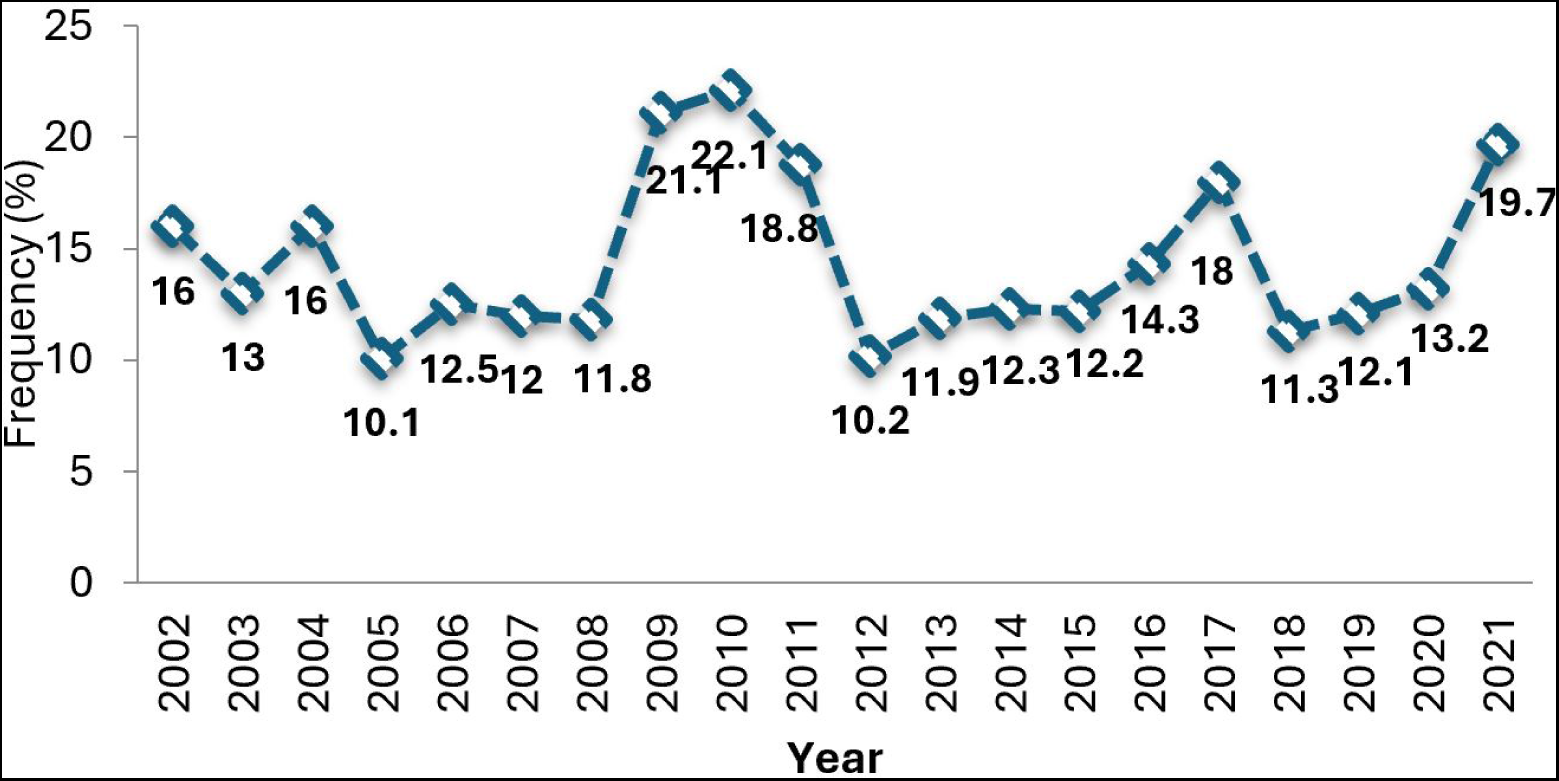
Contraceptive prevalence rate between 2002-2021 Source: Calculations from the Directorate of Programming and Planning (DDP)/Ministry of Health (Benin) data

In Benin, analyses show that the contraceptive prevalence rate is increasing. It went from 16% in 2002 to a high rate of 22.1% in 2010 before falling to 10.2% in 2012. From this date, this rate regained momentum until it reached 18%. in 2017. It fell by 6.7% in 2019 and increased to 19.7% in 2021.

**Figure 17.**
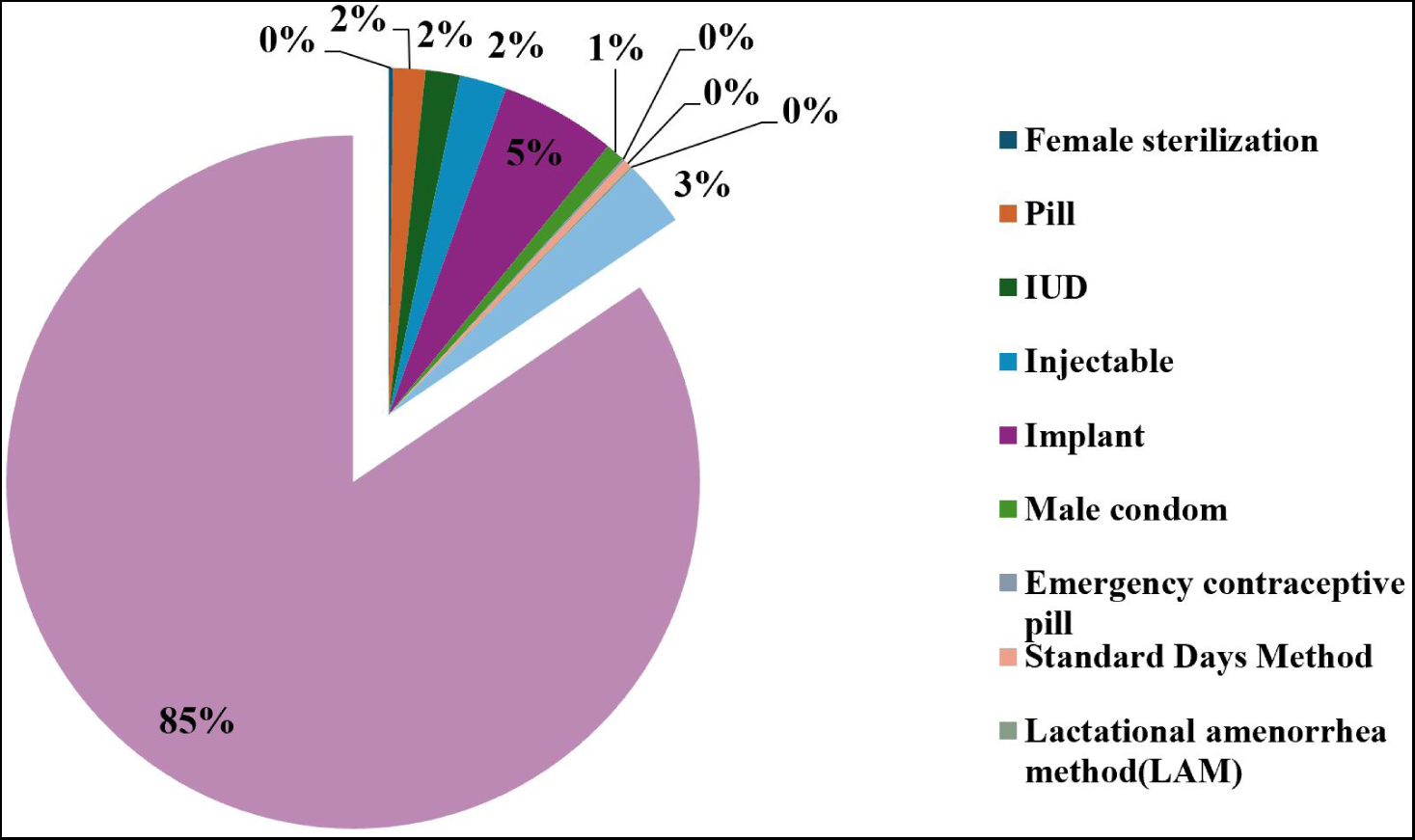
Contraceptive methods used by women in 2017-2018 Source: Calculation from Benin Demographic and Health Survey (DHS) data, 2017-2018

Nearly 85% do not use any contraceptive method. Among those who use them, only 3% practice traditional methods compared to 12% who use modern contraceptive methods (5% implant, 2% pill, 2% IUD, 2% injectable, 1% condom).

## 4 Discussion

Under the terms of this diagnostic study, the analysis of the evolution of the indicators allows us to categorize the progress obtained and the main challenges by area of the health system.

### Financial Resources and Health Spending

In Benin, health occupies a significant part of the general State budget. Estimated at 8.4% in 2005, the share of the State budget devoted to health fell slightly before experiencing a peak of 10.1% in 2009-2010 due to the free cesarean section adopted after diagnosis of level of deaths observed during childbirth and the exorbitant cost of cesarean section. This free cesarean section was one of the greatest social measures in the health sector. From 2011, the drop in the budget allocated to health increased and became considerable, especially after 2016 with numerous reforms carried out by the government to clean up the sector. A breakdown by expenditure item shows that investments in health account for half of expenditure. Personnel expenditure has increased with the increase in the number of staff, the gap of which increases over time due to strong population growth.

### Socio-health infrastructure, health human resources and accessibility to health care

This is where the problem of population growth in Benin finds its meaning. The results indicate a more than doubling increase in unmet needs for healthcare personnel between 2002 and 2021. The number of inhabitants per doctor increased from 7,210 to 13,913 in 20 years. The gap is even greater for the number of inhabitants per nurse, going from 2,440 to 46,778. These gaps have especially worsened with the reform of practice in private clients carried out in 2018 which now requires caregivers to only practice in one single sector (public or private), thus avoiding the duplication of nursing staff which thus veiled the true situation of the country in terms of human resources. This calls into question the efficiency of the use of human resources in the health sector. Normally, strong demographic growth creates more gaps in human resources in terms of health, but the biggest problem is the unequal otherwise poor distribution of these human resources creating inequalities and even frustrations among stakeholders. The bitter observation is that very remote and isolated health centers are rarely without healthcare personnel. This situation will inevitably have negative impacts on the quality of care provided in these settings. On the other hand, there is an improvement in accessibility to health services. Coverage rates increased from 86% in 2002 to 96% in 2021, reflecting the availability of health centers and hospitals for the treatment of patients. Attendance rates at health facilities have also increased from 35% to 56% in 20 years. This is also an effort to be commended.

### Disease prevention and health promotion

A good health system can prevent diseases and implement effective strategies to promote good health. Philippe Geluck’s formula, “Prevention is better than cure”, is well indicated to mean that it is preferable to take measures to avoid diseases as much as possible rather than waiting for them to be monitored before seeking to treat them, especially in a context of limited resources. Treatment should be the last resort. Disease prevention efforts are being implemented in Benin through several key actions and strategies. To better monitor the pregnancies of pregnant women and facilitate a safe delivery, prenatal consultations are organized. The efforts of the Ministry of Health with the support of these partners have made it possible to significantly improve this indicator. The same is true of assisted childbirth where at least one quality staff assists the woman during childbirth unlike previous years (before 2006) where we witnessed many births at home and or by unqualified agents, especially in peripheral health facilities. Progress in the field of vaccination (protects children against certain childhood diseases) is also commendable thanks to the support of partners, mainly UNICEF. Vaccination rates are very encouraging beyond the 100% targets. These efforts have contributed to a significant reduction in infant death rates. Other prevention and health promotion strategies are implemented through family planning programs. The results indicate a slow improvement in contraceptive prevalence rates (11.6% in 2022 to 19.7% in 2021), in sharp contrast to the significant resources being injected into it. This raises the issue of the effectiveness of the communication strategies used. The country would benefit from redirecting the major resources available for planning in other priority sectors, particularly the improvement of the education system capable of producing people because, as the famous Indian economist Amartya Sen said, development is the best contraceptive.

### State of health of the Beninese populations

It is the essential pillar by which we assess the quality and maturity of a health system. Beyond the efforts noted above and the remaining challenges, what can we say about the evolution of the health status of populations? Here a lot of efforts are noted. Despite the increase in the incidence rate of malaria, which still remains the leading cause of consultation, there has been a significant and continuous decline in deaths due to malaria, going from 141 deaths in 2003 to 86 per 100,000 inhabitants in 2021. This decline could explained by the implementation of several strategies to combat malaria with the support of numerous technical and financial partners. These strategies include prevention, case management, advocacy, communication and social mobilization, without forgetting the management of drug supplies and stocks. We also note a significant drop in the prevalence of HIV AIDS going from a stabilization around 1.2% between 2008-2011, to 0.8% from 2019 ahead of several neighboring countries in the sub-region which are for most beyond 1% according to WHO data in 2021. This shows the efforts of the State in the prevention of this endemic disease. The big challenge remains massive screening of the population and putting infected people on antiretrovirals (ARV) to quickly break the chain of contamination in the hope of completely eliminating HIV in Benin. The same goes for the incidence of tuberculosis, whose country’s efforts through the National Tuberculosis Control Program (PNLT) have made it possible to increase from 73 to 53 per 100,000 inhabitants from 2005 to 2020. Deaths and in particular deaths of children under 5 years old have also decreased, thus improving life expectancy from birth from 59.6 years in 2002 to 63.8 years in 2013. Early neonatal mortality has decreased going from 10.1‰ in 2002 to 4.7‰ in 2021. However, this decline remains insufficient because neonatal mortality has been identified as that which contributes to infant deaths. Furthermore, the evolution of maternal deaths since 2018 reveals the insufficiency of strategies to reduce maternal deaths. Everything suggests that the government’s rigor in implementing major reforms in the health field, ceased in 2018 since several indicators declined from this year. The health insurance system in Benin also constitutes a major challenge.

### Health care expenditure and the health insurance mechanism

In Benin, public and private healthcare providers bill the patient who pays directly out of pocket regardless of the amount. For years, the system has been marked by overbidding, especially in private establishments. At the public level, although prices are regulated by determining the average costs of services by pathology and type of care by the model of grouping patients by homogeneous diagnostic groups, we are still witnessing certain bad ones such as ransoms, and overbilling coupled with the sale of parallel drugs. Thus, health care costs constitute a heavy burden for the population, the majority of whom find themselves unable to access quality health care. According to resource persons in the health sector, some of Benin’s workers benefit from health insurance supported by their employers, the health insurance coverage rate of which goes up to 80% for the majority and 100% for a few privileged structures. international organizations. Unfortunately, self-employed workers, traders, actors in the informal system, and certain employees do not benefit from any health insurance and are obliged to pay enormous financial resources to deal with health problems. This is a real social inequality because only a relatively wealthy minority achieves it.

Also, the State has thought about the situation of individuals who have no professional situation to subscribe to compulsory health insurance. For the case of citizens identified as extremely poor and unable to pay their social contributions, the State has set up a special type of insurance. This is the Human Capital Strengthening Insurance (ARCH) financed from the national budget and the contribution of technical and financial partners. This insurance also provides access to a certain number of essential health care services. However, discussions with resource people revealed some difficulties in operationalizing this approach. The populations met mention complaints from stakeholders about not being able to benefit from quality services which will be relevant to evaluate in other work. The state must review the health insurance system by opting for a compulsory system which gives everyone equal access to quality health care, reduces household spending on health, strengthens solidarity and reduces discrimination between subjects. The system must draw its resources from individuals’ contributions in proportion to their income and then pool them to meet health expenses.

### Challenges and perspectives

Despite the efforts and progress recorded by the health system, many challenges remain and deserve particular attention from governments and partners. The country still faces major health problems such as reproductive, maternal, newborn, child, adolescent and elderly health; priority diseases (HIV, Hepatitis, Tuberculosis, malaria, etc.); non-communicable diseases such as cancer, diabetes, cardiovascular diseases, chronic respiratory diseases(World Health Organization, 2021). Furthermore, the cross-analysis of health and socio-economic indicators, added to the strong demographic growth of the country, clearly expresses the threats weighing on the state of health of the Beninese populations and calls for a new way of thinking for a health system. sustainable health.

It will therefore be important to ensure the efficient use of health personnel by ensuring a good distribution of personnel throughout the territory. We must appeal to patriotism through profound reforms of the education system and the introduction of lessons of morality, civics and patriotism into training programs. This will make it possible to improve good practices in the provision of health services and the proper use of available material resources, equipment and socio-sanitary infrastructure. It will also be necessary to strengthen the governance and regulation of the sector to guarantee the accessibility and quality of health services. Great emphasis should be placed on disease prevention and health promotion through effective communication strategies including the promotion of good hygiene and basic sanitation, community communications, and dialogues on health promotion. health, the use of digital channels, and social networks to promote key health messages.

## Acknowledgements

We affirm that this paper is original and is not currently under consideration by any other publication.

## Data availability

The data can be obtained from the corresponding author (on request).

## Ethics Statement

This research does not require ethical approval.

